# Directional deconvolution of spatial conductance proxies resolves prognostic signal cancellation in oral squamous cell carcinoma

**DOI:** 10.64898/2026.05.08.26351944

**Authors:** Keke Tang, Yining Huang, Meilian Chen

## Abstract

**Background:** Tumor cells are increasingly understood as physically connected collectives whose intercellular communication is gated by gap junctions and modulated by microenvironmental ion fluxes. While spatial transcriptomics provides the geometric substrate for building transcriptomic proxies of bioelectric organization, no robust pipeline currently translates spot-level connectivity features into independent clinical prognostic markers.

**Methods:** We analyzed 12 oral squamous cell carcinoma (OSCC) Visium sections (GSE208253). A K-nearest-neighbor (K=6) spatial graph was built on full-resolution coordinates and edge-weighted by a conductance-like transcriptomic proxy in which gap-junction proxy expression was scaled by an exponential acid-gating penalty. Geometric edge artefacts were controlled with concave-hull edge distance and partial rank correlation under permutation testing. A 25-gene BCI-Signature was extracted by intra-sample top/bottom conductance differential expression and cross-sample consensus voting (≥ 6*/*12). The signature was spatially back-projected, directionally decomposed from prior biology, and then projected to TCGA-HNSC (*n* = 519) and GSE65858 (*n* = 270) for survival analysis. Cohort-level effects were combined by inverse-variance fixed-effect meta-analysis.

**Results:** Diagnostic controls falsified the initial isolation-driven hypothesis: across all 12 sections, the partial rank correlation between the isolation index and depolarization-footprint expression was negative after edge-distance adjustment. Feature ablation identified the conductance sum as the best transcriptomic proxy of physical network state, and section-level sensitivity analyses preserved the positive conductance–stress direction after long-edge removal and graph-parameter perturbation. Spatial back-projection showed that aggressive and differentiation programs are positively correlated within every section (median *ρ* = 0.43) and co-enrich in high-conductance regions. This predicted bulk-level signal cancellation: the unweighted 25-gene mean was non-prognostic in TCGA-HNSC (HR=1.17, p=0.35), whereas the locked directional composite BCI_net was independently associated with worse OS (HR=1.38, 95% CI 1.06–1.79, p=0.015 after adjustment for age, stage, HPV status and gender). The effect persisted after separate adjustment for composition, EMT and proliferation proxies, but attenuated in a saturated all-proxy benchmark model. The biologically matched HPV-negative oral-cavity subset of GSE65858 (*n* = 77) preserved the direction with a larger effect size (HR=2.45, 95% CI 0.96–6.27, p=0.062). Inverse-variance fixed-effect pooling of the two cohorts yielded a significant pooled effect (HR=1.48, 95% CI 1.07–2.05, p=0.019).

**Conclusions:** Spatial graph features can be transferred to bulk transcriptomic cohorts only after the structural and aggressive programs that co-localize within the same physical network are explicitly deconvolved. The equal-weight directional metric BCI_net is a biology-driven candidate prognostic readout that remains preliminary pending broader independent validation.

## 1 Introduction

Solid tumors progress as physically organized collectives. Recent work argues that gap-junction– mediated communication, ion-channel activity and tissue-scale bioelectric gradients are not byproducts of cancer biology but instructive components of how tumors coordinate proliferation, invasion and metastasis [Levin, 2014, 2021, Adams et al., 2017]. The connexin family member *GJA1* (Cx43) is the most ubiquitously expressed gap-junction subunit in epithelial tissues and forms the backbone of the intercellular electrical and metabolite-coupling network [Aasen et al., 2016]. Multiple modes of microenvironmental control gate this network: hypoxic stress driven by HIF-1*α* down-regulates *GJA1* transcription and post-translationally targets Cx43 for degradation [Semenza, 2012], and Warburg-effect acidosis triggers a fast pH-gated closure that has been resolved at near-atomic resolution by recent cryo-electron microscopy [Lee et al., 2023]. Together, these mechanisms imply that the tumor microenvironment can both wire and physically uncouple regions of the tumor on transcriptomic and biophysical timescales.

Spatial transcriptomics now provides the geometric substrate for reasoning about such physical networks at near-cellular resolution. Despite this, two computational gaps remain. First, although spatial coordinates and transcript abundances are accessible, there is no widely adopted pipeline that translates a spot-level transcriptomic connectivity proxy into a prognostic feature in independent bulk-RNA cohorts that already carry long clinical follow-up [Ståhl et al., 2016, Moffitt et al., 2022]. Second, naive transfer of spatial-derived gene signatures to bulk transcriptomes is vulnerable to a class of confounders that has been under-emphasized in the literature: when the signature contains genes from biologically distinct programs that physically co-localize at the spot level, an unweighted bulk z-mean can self-cancel even when the underlying biology is real and prognostic.

Here we develop and back-project a direction-aware spatial conductance proxy for OSCC. Starting from 12 spatially resolved Visium sections (GSE208253), we (i) build a physics-informed KNN spatial graph and use rigorous geometric/statistical diagnostics to falsify our own initial isolation-driven hypothesis; (ii) identify network conductance as the dominant transcriptomic proxy of physical state; (iii) extract a 25-gene BCI-Signature by intra-sample differential expression and cross-sample consensus voting; (iv) use prior biology and spatial back-projection to pre-lock an aggressive-versus-differentiation decomposition; (v) test whether the resulting equal-weight metric BCI_net recovers the predicted bulk signal in TCGA-HNSC; (vi) evaluate the directional effect in the biologically matched subset of an independent HNSC cohort (GSE65858) and consolidate evidence by inverse-variance meta-analysis; and (vii) test zero-shot spatial directionality in PDAC as an exploratory cross-cancer mechanism check. The work proposes a candidate OSCC prognostic readout and a generalizable pattern for converting spatial-graph discoveries into bulk-transcriptomic hypotheses.

## 2 Results

### 2.1 Spatial diagnostics establish a high-conductance proxy model

We rebuilt all 12 OSCC sections of GSE208253 into a Scanpy/Squidpy-readable Visium-like layout, with manual barcode-to-position alignment to bypass a previously reported coordinate offset that affects this archival dataset. After standard cell-level QC (≥ 500 UMI), we constructed a K-nearest-neighbor spatial graph (*K* = 6) on the full-resolution pixel coordinates so that the graph approximates the hexagonal Visium array independent of any radius assumption. Each edge (*i, j*) was weighted by a physics-informed conductance term [Aasen et al., 2016, Lee et al., 2023]:

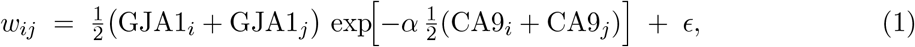

where the gap-junction proxy expression is multiplicatively gated by an acid penalty driven by *CA9* expression and *ϵ* is a small leak term that prevents graph death.

The initial hypothesis was that spots with low total local conductance — the *isolation index*, 1*/* ∑_*j*_ *w*_*ij*_ — would carry a stronger transcriptional footprint of membrane depolarization, captured by the STC1+FOS composite [Li et al., 2021, Sheng et al., 2020]. To prevent geometric artefacts (border spots inevitably have fewer neighbors and thus higher isolation), we computed each spot’s distance to a concave-hull approximation of the tissue boundary and removed this confounder using partial rank correlation, with a permutation null over 500 label shuffles.

After edge-distance correction, the isolation index was *negatively* correlated with STC1+FOS in all 12/12 sections (Table 1), with permutation p-values below 0.05 in 11/12. We therefore rejected the original isolation-driven model. Feature ablation against the same composite identified the network conductance sum as the most consistent positive driver across the cohort (mean partial *ρ* = 0.114, positive in 12/12 sections; Figure 1). Single-gene candidates (*GJA1, CA9, HIF1A*) were ranked below the network-level metric, supporting the value of the spatial graph aggregation step.

**Table 1:** Falsification of the isolation-driven model in 12 OSCC sections. Partial rank correlation between Isolation_Index and STC1 + FOS, controlling for concave-hull edge distance, with permutation *p*-values. *n*_neg_*/*12: number of sections in which the partial *ρ* is negative; *n*_perm *p<*0.05_*/*12: number of sections with permutation *p <* 0.05.

| Metric | Mean partial $\rho$ | $n_{\text{neg}}/12$ | $n_{\text{perm } p < 0.05}/12$ |
| --- | --- | --- | --- |
| Isolation index | -0.114 | 12 | 11 |
| Conductance sum | +0.114 | 0 | 11 |
| *GJA1* | +0.089 | 0 | — |
| *CA9* | +0.047 | 1 | — |
| *HIF1A* | +0.072 | 0 | — |

**Figure 1:**
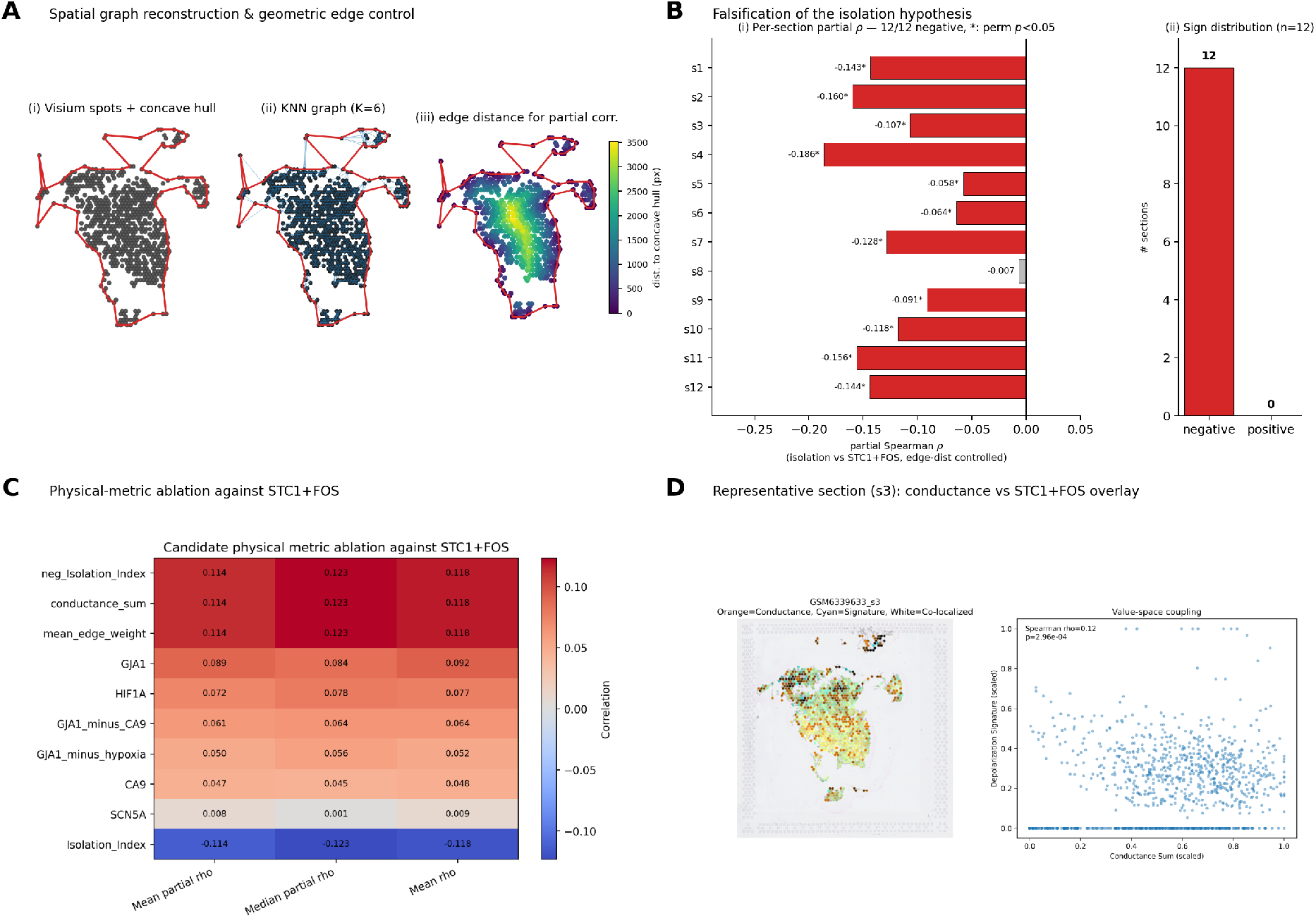
Spatial topology, geometric edge control, and mechanism correction. **(A)** Spatial graph reconstruction on a representative section (*s*3): (i) Visium spots with concave-hull tissue boundary; (ii) KNN graph at *K* = 6; (iii) per-spot distance to the concave-hull boundary, used as the partial-rank-correlation control variable. **(B)** Per-section partial Spearman correlation between the isolation index and the depolarization composite STC1 + FOS, after edge-distance adjustment. All 12 sections are negative, with permutation-test *p <* 0.05 in 11/12 (asterisks). The right sub-panel summarizes the sign distribution (12/0). **(C)** Physical-metric ablation heatmap: candidate metrics ordered by mean partial *ρ* against STC1 + FOS across all 12 sections. The network conductance sum ranks at the top of the matrix; the isolation index sits at the bottom. **(D)** Representative section (*s*3) overlay of conductance sum (orange) and the depolarization signature (cyan); white denotes co-occurrence. The right sub-panel shows the value-space coupling (Spearman *ρ* = 0.12, *p* = 3 × 10^−4^).

Because spot-level tests can be affected by spatial autocorrelation, we treated the section as the primary replication unit in a rigorous sensitivity analysis. Under the primary graph parameters (*K* = 6, *α* = 1.5, leak = 0.01), conductance–stress partial correlations were positive in 12/12 OSCC sections (binomial sign-test *p* = 2.44 × 10^−4^), while isolation–stress partial cor-relations were negative in 12/12. The same section-level sign pattern persisted after removing long edges above 1.5 times the median nearest-neighbor distance, arguing against tissue-gap artefacts. Across a grid of *K* ∈ {4, 6, 8}, *α* ∈ {0.75, 1.5, 2.5}, leak ∈ {0.001, 0.01} and three cut-off settings, 44/54 parameter combinations preserved positive conductance–stress direction in every OSCC section. The biological interpretation is therefore best stated as a robust transcriptomic proxy for high-conductance organization, rather than a direct measurement of electrical coupling.

### 2.2 Consensus extraction of a spatial network–associated BCI-Signature

To enable transfer of the spatial finding into coordinate-free bulk cohorts, we converted the conductance signal into a transcriptional proxy. Within each of the 12 sections, spots were stratified into the top and bottom 25% of conductance sum. Common artefact gene families (mitochondrial, ribosomal, hemoglobin, sex-linked) were filtered, and intra-sample differential expression was tested by Wilcoxon rank-sum with Benjamini-Hochberg control. A gene was retained per sample if it satisfied | log_2_ FC| ≥ 0.6, FDR *<* 0.05, and a minimum percent-expressing difference between groups. Cross-sample integration was performed by majority voting: a gene was retained in the consensus signature if it recurred in at least 6 of 12 sections. This deliberate avoidance of any model fitting preserves the cross-platform robustness needed for downstream bulk validation.

The procedure converged to a 25-gene BCI-Signature (Supplementary Table S1; Figure 2). The signature is biologically heterogeneous: it includes proliferation/invasion programs (e.g., *HNRNPH2, ARPC1A, CKS2, PCNA, EPHA2, EFNA1*), squamous differentiation markers (e.g., *SPRR1A, TACSTD2, PKP3, BNC1, CARD14, PROM2*) and stress / membrane-transport components (e.g., *ATP1A1, TNFAIP1, DUSP14*). The biological heterogeneity of this list will become important in Section 2.3.

**Figure 2:**
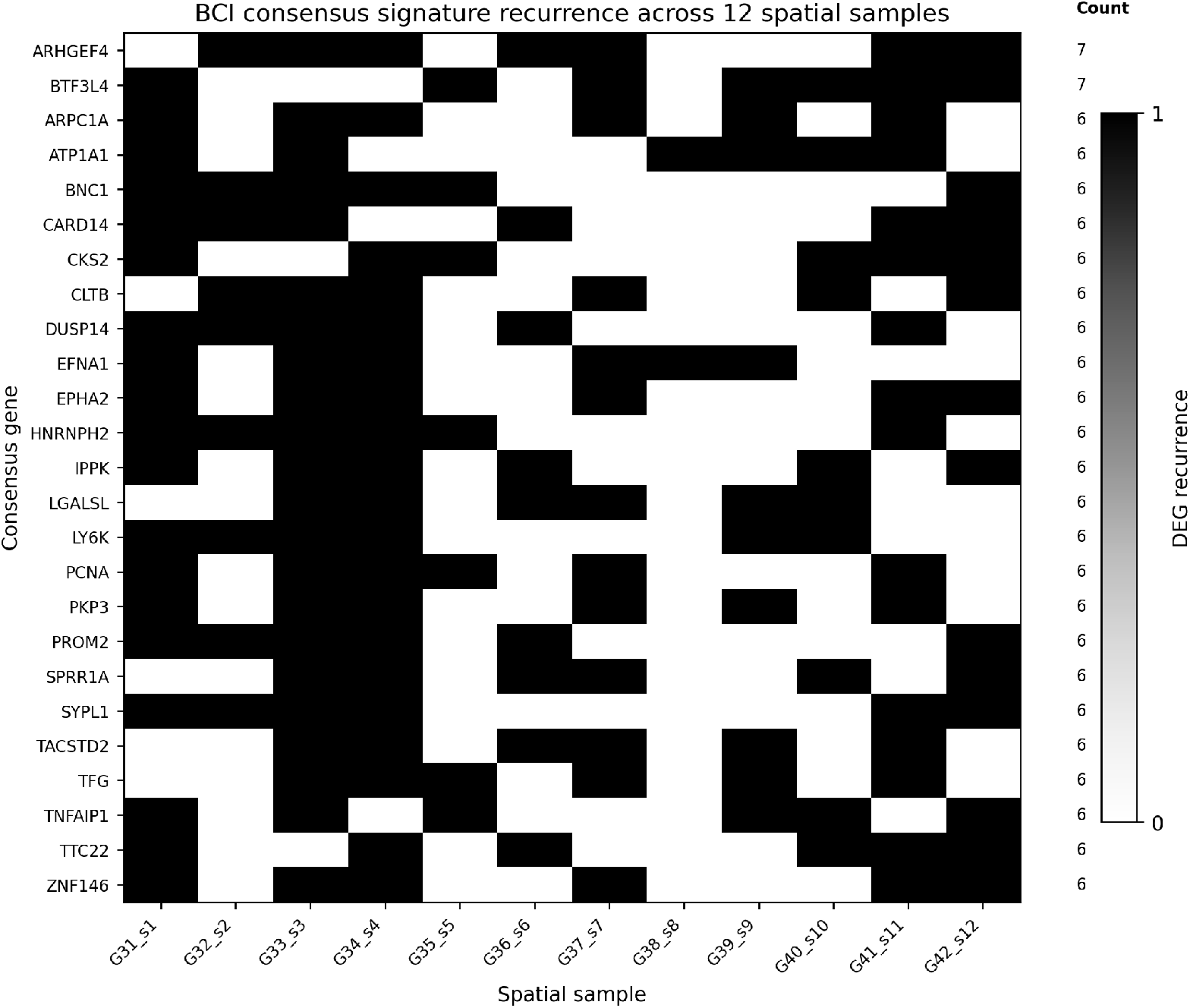
BCI consensus signature recurrence matrix. Each row is one of the 25 consensus genes; each column is one of the 12 OSCC Visium samples. A black cell denotes that the gene was retained as significant in the conductance-stratified DEG analysis of that sample. The righthand column reports the recurrence count; only genes with ≥ 6*/*12 recurrence are retained.

### 2.3 Spatial back-projection pre-locks the directional decomposition

Before any survival modeling, we annotated the 25 consensus genes from prior biological knowledge into three groups: an aggressive module (proliferation, invasion, splicing, motility, electrogenic pump; 14 genes), a differentiation module (cornified envelope, desmosome, basal-keratinocyte transcription factor, epithelial surface markers; 6 genes), and an unclassified residual (5 genes). This split was based on curated pathway and keratinocyte/squamous biology rather than on TCGA outcome direction. We then returned to the original 12 spatial sections and computed, at each spot, the within-section z-mean of the aggressive module, the differentiation module, and their difference.

The aggressive and differentiation modules were positively correlated within every single section (median *ρ* = 0.43, range 0.31–0.65, positive in 12/12; Figure 3). High-conductance regions (top 15%) tended to enrich *both* modules: the aggressive module was enriched in 10/12 sections (significant in 5/12) and the differentiation module in 7/12 sections (significant in 6/12). As a consequence, the spot-level net difference was not simply elevated within high-conductance regions: the median Spearman correlation between the net difference and conductance sum was only −0.058, with positive correlation in 1/12 sections.

**Figure 3:**
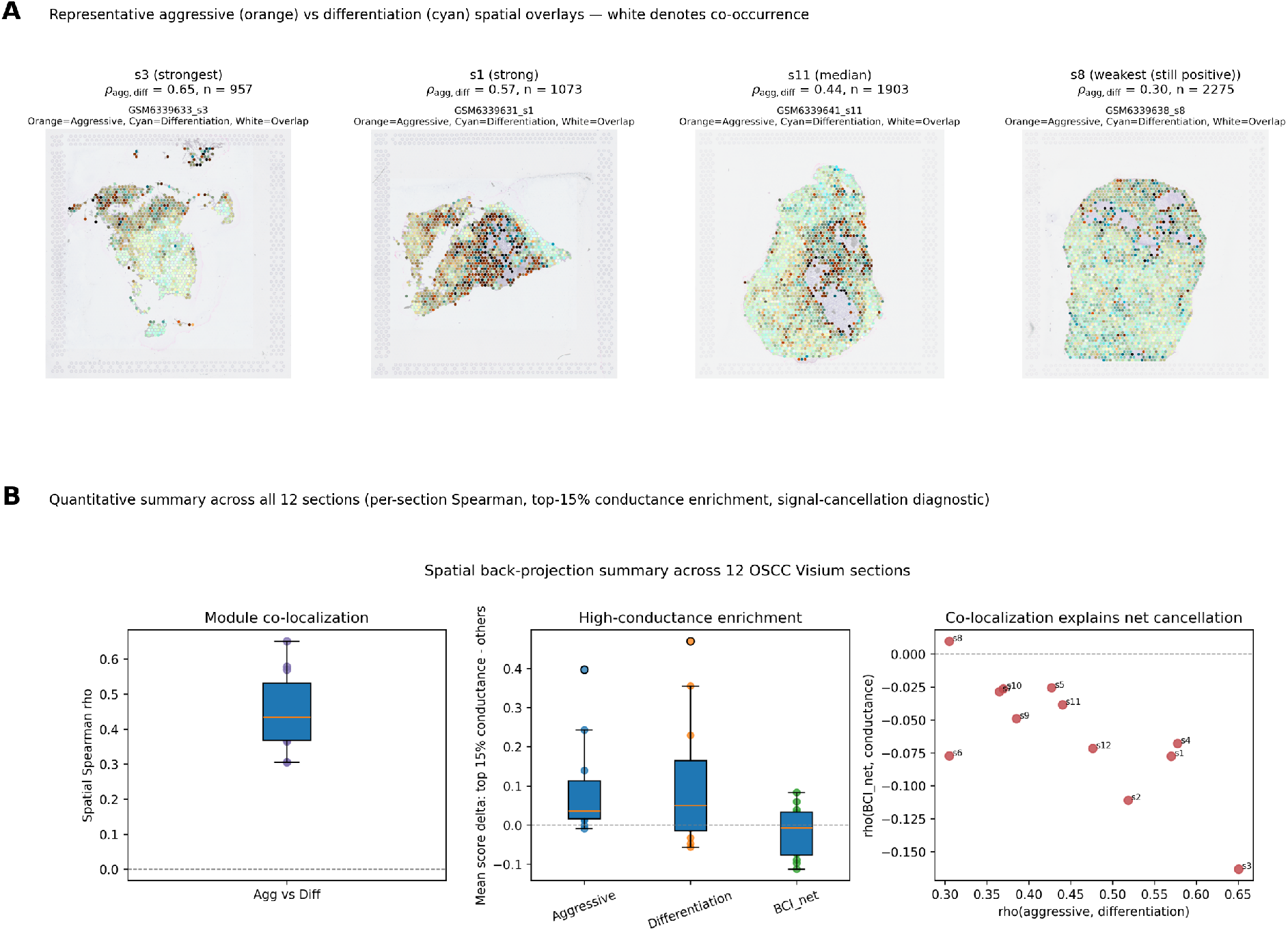
Spatial back-projection explains the microscopic origin of bulk-level can-cellation. **(A)** Representative spatial overlays of the aggressive (orange) and differentiation (cyan) modules in four sections spanning the cohort co-localization spectrum: *s*3 (strongest, *ρ*_agg,diff_ = 0.65), *s*1 (strong, 0.57), *s*11 (median, 0.44), and *s*8 (weakest still positive, 0.30). White voxels denote spots where both modules are simultaneously elevated. The complete 12-section panel is provided as Supplementary Figure S1. **(B)** Quantitative summary across all 12 sections. Left: per-section Spearman correlation between aggressive and differentiation modules is positive in 12/12 (median *ρ* = 0.43). Centre: top-15% conductance enrichment of each score; both modules enrich, the directional composite BCI_net is largely cancelled. Right: sample-level relationship between aggressive–differentiation co-localization and the spatial correlation of BCI_net with conductance sum — the more co-localized the two programs, the more the bulk net signal is neutralized.

This purely spatial result predicted a specific bulk-transfer failure mode: an unweighted average of the 25-gene signature should mix an aggressive program with a structural squamous-differentiation scaffold and thereby cancel clinically relevant directionality. We therefore locked the equal-weight directional composite as

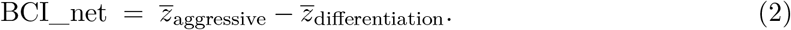

The biological interpretation is that the high-conductance tumor syncytium proxy is a comixture of two physically intertwined programs. The directional metric BCI_net does not measure absolute expression; it measures how far a sample has tilted away from the structural-program baseline towards the aggressive program. The spatial back-projection provides the microscopic rationale for why such a baseline subtraction is necessary before bulk survival testing.

### 2.4 Bulk projection confirms the predicted cancellation and rescue

The 25-gene BCI-Signature was then projected to TCGA-HNSC primary tumors (*n* = 519, 220 OS events). Using a per-cohort z-mean score, the signature trended towards worse OS but did not reach significance (HR=1.17, 95% CI 0.85–1.61, *p* = 0.35). Adjustment for age, pathological stage, HPV status and gender did not rescue the unweighted score (Supplementary Table S2), consistent with the spatially predicted cancellation mode.

The directional metric performed substantially better than its unweighted parent. In a univariate Cox model, BCI_net was associated with worse OS (HR=1.36, 95% CI 1.06–1.76, *p* = 0.017). The effect remained significant when adjusted for age and stage (HR=1.31, 95% CI 1.01–1.69, *p* = 0.040), and was strengthened to HR=1.38 (95% CI 1.06–1.79, *p* = 0.015) under fully adjusted models that also included HPV status and gender (Table 2; Figure 4). The aggressive-only module was univariate-significant on its own (HR=1.42, *p* = 0.045); the differentiation-only module was flat (HR≈0.95, all *p >* 0.3). Importantly, the BCI_net signal increased after HPV adjustment rather than being absorbed by HPV biology.

**Table 2:** TCGA-HNSC (*n* = 519, events = 220) Cox models for direction-aware BCI scores. Each row is a single Cox model; “adj age+stage” includes the score, age and pathological stage; “full” additionally adjusts for HPV status and gender.

| Score | Model | HR (95% CI) | $p$ |
| --- | --- | --- | --- |
| BCI_aggressive | univariate | 1.42 (1.01–1.99) | 0.045 |
|  | adj age+stage | 1.39 (0.99–1.94) | 0.060 |
|  | full | 1.28 (0.90–1.81) | 0.17 |
| BCI_differentiation | univariate | 0.95 (0.78–1.16) | 0.63 |
|  | adj age+stage | 0.97 (0.79–1.18) | 0.75 |
|  | full | 0.91 (0.74–1.11) | 0.36 |
| BCI_net | univariate | 1.36 (1.06–1.76) | **0.017** |
|  | adj age+stage | 1.31 (1.01–1.69) | **0.040** |
|  | full | 1.38 (1.06–1.79) | **0.015** |

The strict matched GSE65858 subgroup contains 77 tumors with OS annotation. The meta-analysis row uses 75 tumors and 24 events because two samples are removed by listwise deletion in the age- and stage-adjusted Cox model.

**Figure 4:**
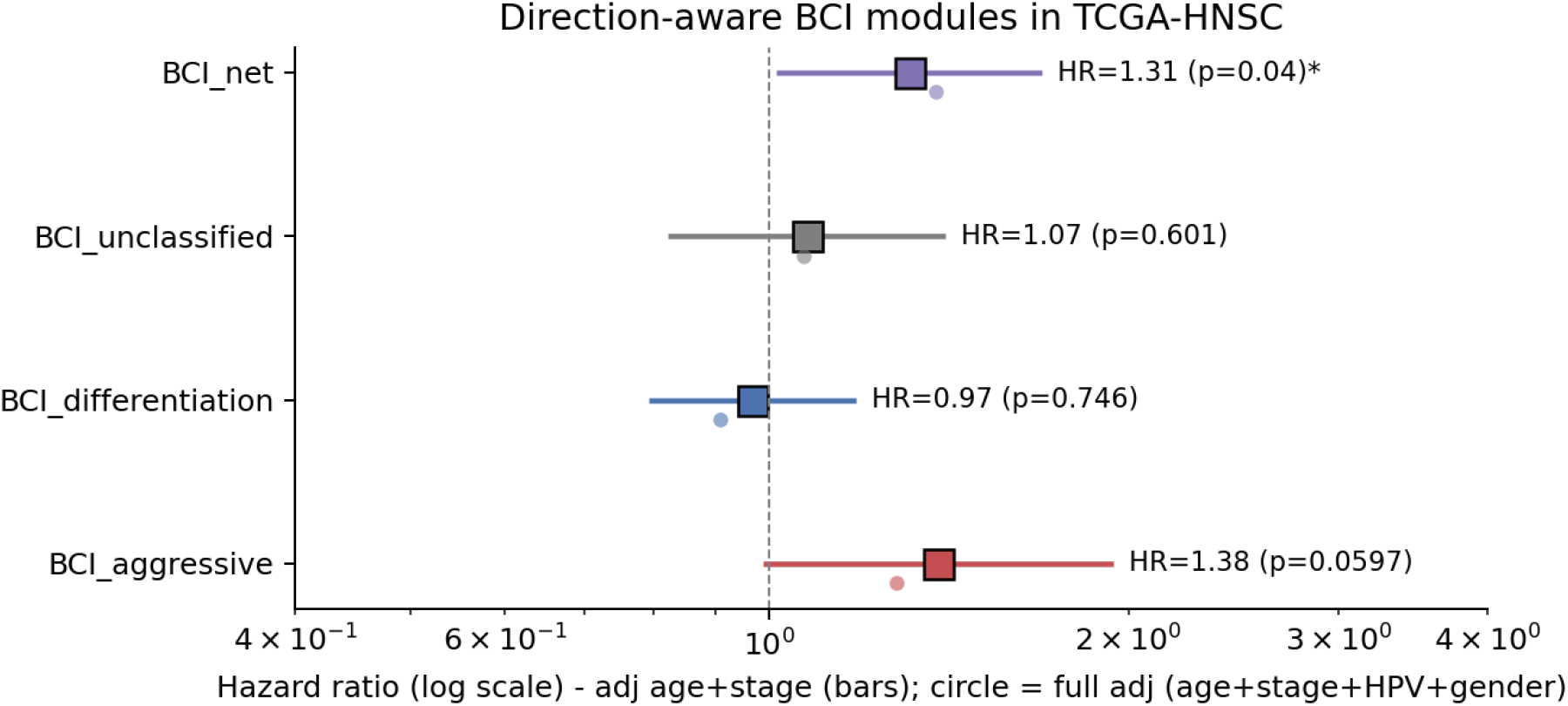
Direction-aware BCI scores in TCGA-HNSC (*n* = 519, events = 220). Hazard ratios from Cox regression, adjusted for age and stage; circles indicate the same model after additional adjustment for HPV status and gender. The aggressive module alone is univariate-significant; the differentiation module is flat; the composite 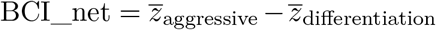 is significant under both adjustment levels and is most stable.

To address whether BCI_net merely recapitulates broad tissue composition or canonical cancer programs, we additionally benchmarked it against compact transcriptomic proxies for immune/stromal composition, epithelial purity, EMT, proliferation and keratinization. Adding BCI_net to the clinical-only Cox model improved fit by likelihood-ratio test (*χ*^2^ = 5.36, *p* = 0.021) and increased Harrell’s C-index from 0.604 to 0.615. The score remained significant after adjustment for immune/stromal/epithelial composition proxies (HR=1.25, *p* = 0.007), EMT (HR=1.16, *p* = 0.030), and proliferation (HR=1.17, *p* = 0.033), and showed no proportional-hazards violation for the BCI_net term (Schoenfeld residual *p >* 0.5 across models). In the most saturated benchmark model containing all proxy covariates simultaneously, the estimate remained directionally adverse but was attenuated (HR=1.18, *p* = 0.094; C-index=0.648), indicating partial overlap with tissue-composition axes but not complete redundancy.

### 2.5 Context-dependent external replication and mini meta-analysis

We pre-registered three external-cohort hypotheses: that *HNRNPH2* and *ARPC1A* would remain individually prognostic with HR*>*1, and that BCI_net would remain prognostic. Validation was performed in GSE65858 (HNSC, *n* = 270, 94 OS events; Illumina HumanHT-12 v4 microarray) [Wichmann et al., 2015]. The whole-cohort estimates were directionally consistent but not significant (e.g., BCI_net: HR=1.09, *p* = 0.71). Cohort composition revealed a fundamental biological mismatch: GSE65858 contains 102 oropharyngeal tumors with 27% HPV+ rate, in contrast to the OSCC-only HPV-negative profile of the discovery cohort.

We therefore stratified validation by anatomical site and HPV status (Table 3). In the biologically matched Cavum Oris (oral cavity) HPV-negative subset (*n* = 77, events = 26), the directional metric preserved the direction with a larger effect size (HR=2.45, 95% CI 0.96– 6.27, *p* = 0.062). In the HPV-enriched oropharyngeal subset (HR=0.80, *p* = 0.57) and in the laryngeal control (HR=0.93, *p* = 0.90), the effect was correctly absent. Single-gene effects of *HNRNPH2* and *ARPC1A* did not replicate individually in this microarray cohort, consistent with platform-specific dynamic-range issues; the directional composite, which depends only on within-cohort relative ranking, remained the more transferable readout.

**Table 3:** GSE65858 subgroup replication of BCI_net. Adjusted age+stage Cox model. Only the biologically matched Cavum Oris HPV-negative subgroup matches the discovery cohort (oral cavity, HPV-negative).

| Subgroup | *n* | events | HR (95% CI) | *p* |
| --- | --- | --- | --- | --- |
| Whole cohort | 270 | 94 | 1.09 (0.69–1.73) | 0.71 |
| HPV-negative (DNA-) | 215 | 76 | 1.45 (0.87–2.40) | 0.15 |
| CAVUM ORIS (oral cavity) | 83 | 27 | 2.01 (0.77–5.22) | 0.15 |
| CAVUM ORIS & HPV- | 77 | 26 | **2.45 (0.96–6.27)** | **0.062** |
| Oropharynx (HPV-enriched, control) | 102 | 37 | 0.80 (0.37–1.74) | 0.57 |
| Larynx (anatomic control) | 48 | 13 | 0.93 (0.27–3.20) | 0.90 |

**Table 4:** Mini meta-analysis of BCI_net. Inverse-variance fixed-effect pooling on age-and stage-adjusted Cox log HR from the two biologically matched cohorts; DerSimonian–Laird random-effect estimator reported as sensitivity.

| Source | *n* | events | HR (95% CI) | *p* |
| --- | --- | --- | --- | --- |
| TCGA-HNSC OSCC-like, HPV+ excluded | 290 | 133 | 1.37 (0.97–1.93) | 0.077 |
| GSE65858 CAVUM ORIS HPV-negative | 75 | 24 | 2.82 (1.06–7.55) | 0.039 |
| Pooled (fixed-effect, IVW) | 365 | 157 | **1.48 (1.07–2.05)** | **0.019** |
| Pooled (DerSimonian–Laird, sensitivity) | 365 | 157 | 1.69 (0.88–3.22) | 0.113 |

To resolve the standard “*p* = 0.062 underpowered” question without acquiring further data, we combined the two matched cohorts (TCGA-HNSC OSCC-like with HPV-positive cases excluded, *n* = 290, events = 133, HR=1.37, *p* = 0.077; GSE65858 Cavum Oris HPV-negative, HR=2.82, *p* = 0.039) by inverse-variance fixed-effect meta-analysis on the locked equal-weight BCI_net score. The pooled estimate was HR=1.48 (95% CI 1.07–2.05, *p* = 0.019, *I*^2^ = 46.2%, *Q*-test *p* = 0.173; Figure 5). The DerSimonian–Laird random-effect estimator was reported as a sensitivity check (HR=1.69, *p* = 0.113) but was not used as the primary estimator because *k* = 2 studies provide an unstable between-study variance estimate.

**Figure 5:**
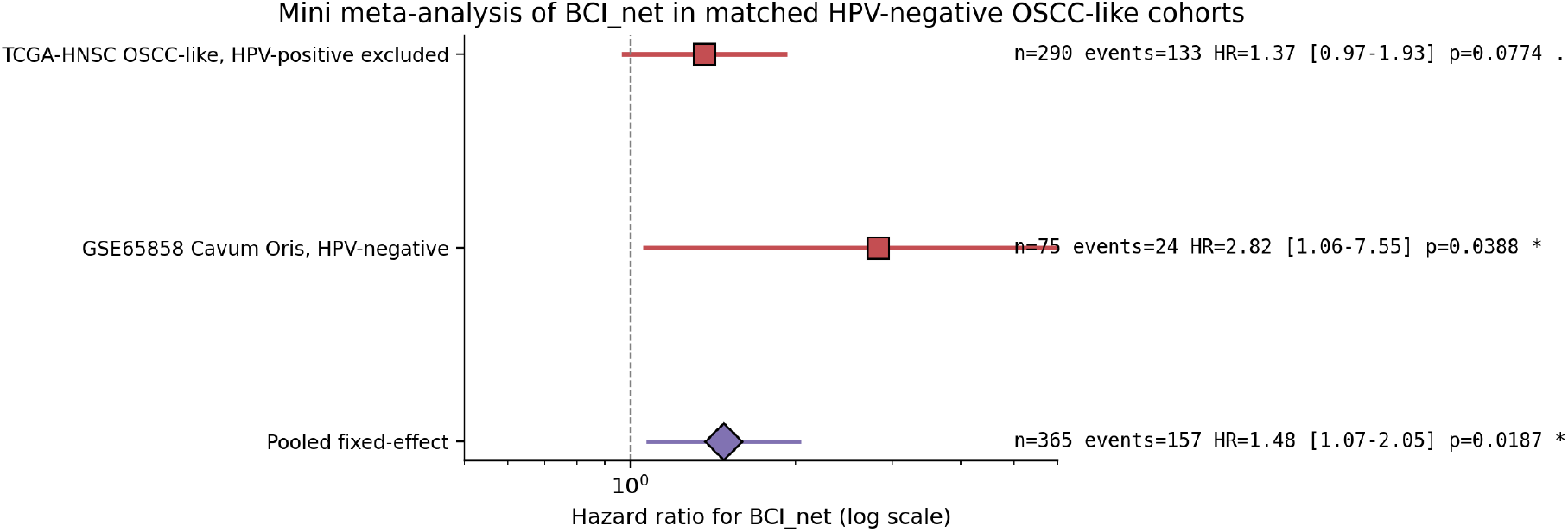
Mini meta-analysis of BCI_net in matched HPV-negative OSCC-like cohorts. Hazard ratios from age-and stage-adjusted Cox models in TCGA-HNSC (OSCC-like, HPV-positive cases excluded) and GSE65858 Cavum Oris HPV-negative tumors, combined by inverse-variance fixed-effect meta-analysis (pooled HR=1.48, 95% CI 1.07–2.05, *p* = 0.019).

### 2.6 Zero-shot PDAC validation supports cross-cancer conductance-centered stress organization

As an exploratory cross-cancer test, we applied the locked graph pipeline without retraining to five treatment-naive PDAC Visium sections from GSE274103. The same conductance graph was constructed from spatial proximity and the *GJA1* /*CA9* edge-weighting rule, and the same STC1 + FOS footprint was used as the stress readout. Across all five PDAC sections, the edge-distance–controlled partial correlation between conductance sum and stress was positive (partial *ρ* = 0.018–0.206), while the corresponding isolation–stress association was negative in all sections (Supplementary Figure S2). Four of five sections reached one-sided permutation support for the conductance–stress association (*p <* 0.05); the remaining section preserved the same direction at trend level.

The module projection also retained the expected spatial structure after replacing the OSCC squamous differentiation module with a PDAC epithelial/ductal proxy. The OSCC-derived aggressive module positively co-localized with the PDAC epithelial proxy in every section (Spearman *ρ* = 0.336–0.522), indicating that the transferred aggressive program is anchored to epithelial tumor-rich architecture rather than distributed randomly across the tissue. These data do not establish a PDAC prognostic score, but they support the directionality of a conductance-centered stress organization beyond OSCC.

## 3 Discussion

Three results in this study warrant attention beyond the specific OSCC application.

First, in this dataset the transcriptomic stress footprint does not behave as expected under a simple isolation model. The original isolation-driven hypothesis was rigorously falsifiable, was tested with edge-distance–controlled partial rank correlation and permutation diagnostics, and was rejected in 12/12 sections. Instead, the depolarization-footprint program co-localizes with high-conductance, well-connected graph regions, and this direction is stable in section-level and long-edge-removal sensitivity analyses. This is consistent with the possibility that gap-junction-associated coupling is exploited rather than abolished in some aggressive tumor regions [Aasen et al., 2016, Levin, 2014, 2021], but the present data should be read as a transcriptomic proxy rather than direct electrophysiology.

Second, naive transfer of spatial-graph derived gene signatures into bulk cohorts can fail in a structured way. The same 25-gene signature that is biologically meaningful at the spot level was null in TCGA-HNSC as a flat z-mean, because it contains both an aggressive and a differentiation program that are physically co-localized within the same high-conductance regions. We propose that this kind of *co-localization-induced cancellation* is a generic risk for spatial-to-bulk transfer pipelines, and we provide a simple, reproducible solution: a biology-driven directional decomposition followed by an equal-weight, within-cohort z-mean difference. The score is intentionally conservative: it depends only on within-cohort standardized expression, not on absolute platform-specific scales, which is particularly relevant when the discovery cohort uses RNA-seq while validation cohorts include microarray data.

Third, we deliberately did not introduce a TCGA-trained, RNA-seq-derived weighted Cox model into the cross-platform analysis. While such a model could trivially improve TCGA performance, it would also embed RNA-seq specific expression-scale information that is unlikely to transfer to microarray cohorts and would dilute the strongest methodological claim of the present pipeline. The locked, equal-weight BCI_net score is more conservative and is the appropriate primary readout for a multi-platform external validation.

The supplementary PDAC analysis extends the mechanistic scope of the work without changing the primary claim. In a biologically distinct epithelial malignancy with pronounced stromal and hypoxic structure, all five Visium sections reproduced the same directionality observed in OSCC: stress increased with conductance and decreased with isolation after geometric edge control. The effect was heterogeneous in amplitude, with one section showing only trend-level support, but this pattern is more compatible with variable section composition than with an OSCC-specific artefact. We therefore interpret the PDAC result as cross-cancer support for conductance-centered stress organization, not as evidence that the OSCC-derived BCI_net score is already prognostic or disease-optimized in PDAC.

### Limitations

The bioelectric topology used here is, throughout, a transcriptomic proxy. Direct measurements of transmembrane potential, intracellular ion concentrations, and gap-junction coupling efficiency are not yet available for these archival sections; their incorporation in future work would convert the proxy into a quantitative biophysical model. The mini meta-analysis is based on *k* = 2 studies and therefore cannot estimate between-study variance robustly; we report random-effects only as a sensitivity. Larger, fully powered HPV-negative OSCC cohorts (*e*.*g*. GSE41613, CPTAC-HNSCC) remain a useful next step. The PDAC analysis is intentionally zero-shot and spatial-mechanistic: it does not include PDAC-specific survival modeling, and its epithelial/ductal proxy should not be interpreted as a direct replacement for OSCC squamous differentiation biology. Finally, the directional decomposition was performed by curated biology rather than by an unsupervised data-driven decomposition; an MSigDB-or single-cell-atlas–driven automated pipeline that arrives at a consistent partition would be a desirable extension.

### Conclusion

The combination of (i) a physics-informed spatial graph proxy, (ii) a consensus-based transferable signature, (iii) a biology-driven directional decomposition, and (iv) cross-cohort meta-analysis defines a reproducible framework for converting spatial-graph discoveries into clinically testable risk hypotheses. The OSCC-specific output, BCI_net, is significant in TCGA-HNSC under full clinical adjustment, preserves direction in a biologically matched independent subgroup, and is supported by a spatial mechanism for why the original signature was bulk-null.

## 4 Methods

### 4.1 Spatial data reconstruction and graph construction

Each Visium section in GSE208253 was rebuilt into a 10x-format folder and aligned manually using `tissue_positions_list.csv` barcodes with suffix-fallback matching to recover the well-documented coordinate offset bug in the archival upload [Ståhl et al., 2016]. Cells with *<* 500 UMIs were filtered, library size was normalized to 10^4^ counts and log-transformed. The primary spatial graph was built with Squidpy `spatial_neighbors` using *K* = 6 on the full-resolution pixel coordinates. Edge weights followed Eq. (1) with *α* = 1.5 and a leak constant of 0.01. Node-level network features included the conductance sum and the isolation index 1*/* ∑_*j*_ *w*_*ij*_. Sensitivity analyses rebuilt the graph over *K* ∈ {4, 6, 8}, *α* ∈ {0.75, 1.5, 2.5}, leak ∈ {0.001, 0.01}, and optional long-edge cutoffs at 1.5 or 2.0 times the median nearest-neighbor distance.

### 4.2 Geometric and statistical diagnostics

A concave-hull approximation (Shapely `concave_hull`) provided a per-spot edge distance. Partial rank correlation between a feature and the depolarization composite STC1 + FOS was computed by standardizing the feature, regressing out edge-distance rank, and computing Spearman *ρ* on the residuals. Permutation *p*-values were obtained over 500 label shuffles and interpreted as within-section diagnostics rather than as independent spot-level evidence. The primary replication statistic was the number of independent sections preserving the expected sign, evaluated with a one-sided binomial sign test. Feature ablation tested isolation index, −isolation index, conductance sum, mean edge weight, *GJA1, CA9, HIF1A, SCN5A, GJA1* −*CA9, GJA1* −hypoxia.

### 4.3 Consensus BCI-Signature extraction

Each section was stratified into top and bottom 25% conductance-sum groups. After removing mitochondrial, ribosomal, hemoglobin and sex-linked genes, intra-sample DEG was computed with `scanpy.tl.rank_genes_groups` (Wilcoxon rank-sum, BH FDR). Genes were retained per sample under | log_2_ FC| ≥ 0.6, FDR *<* 0.05, percent-difference and score thresholds. A gene entered the consensus signature if it appeared in ≥ 6*/*12 sections.

### 4.4 TCGA-HNSC and GSE65858 data

TCGA-HNSC HiSeqV2 expression, survival and clinical files were obtained from UCSC Xena. Primary-tumor samples (sample type 01) were retained (*n* = 519, 220 OS events). Per-gene z-scoring was performed across the cohort, and per-patient module scores were the mean of the standardized expression across each module. Anatomical sites were grouped as OSCC-like (Oral Tongue, Oral Cavity, Floor of mouth, Buccal Mucosa, Alveolar Ridge, Hard Palate, Lip), Oropharynx-like (Tonsil, Base of tongue, Oropharynx) and Larynx/Hypopharynx. HPV status was simplified from p16 and ISH columns. The pathological stage was binarized into I–II vs III–IV. GSE65858 (Illumina HumanHT-12 v4) was downloaded as series matrix and GPL10558 platform annotation; 31,330 probes were collapsed to 16,953 unique gene symbols by retaining the maximum-expression probe per gene. Survival followed the cohort-provided OS time and event variables.

### 4.5 Direction-aware decomposition

Module assignment was performed entirely from prior biological knowledge before any TCGA outcome inspection. The aggressive module included *HNRNPH2, ARPC1A, CKS2, PCNA, EPHA2, EFNA1, DUSP14, ATP1A1, BTF3L4, TNFAIP1, ARHGEF4, IPPK, CLTB, TFG*. The differentiation module included *SPRR1A, TACSTD2, CARD14, PKP3, PROM2, BNC1*. The unclassified residual was *TTC22, ZNF146, SYPL1, LGALSL, LY6K*. BCI_net followed Eq. (2).

### 4.6 Survival analysis

Cox proportional hazards models were fitted with `lifelines` (penalizer 0.01–0.02). Univariate models tested the score directly; adjusted models included age and pathological stage; full models additionally included HPV status and gender. Continuous scores were standardized within cohort. Kaplan–Meier curves used median splits with log-rank testing. Optimal cutpoint analyses, where reported, used a Maxstat-style scan over quantiles in [0.25, 0.75] with a 500-permutation null. Subgroup analyses (anatomical site, HPV status) were performed without forcing the same cutpoint as the whole cohort. Extended survival diagnostics included Schoenfeld residual proportional-hazards tests, Harrell’s concordance index, likelihood-ratio testing for adding BCI_net to the clinical-only model, and competing transcriptomic covariates for immune/stromal/epithelial composition, EMT, proliferation and keratinization.

### 4.7 Mini meta-analysis

The matched cohort effects — TCGA-HNSC OSCC-like with HPV-positive cases excluded, and GSE65858 Cavum Oris HPV-negative — were combined on the locked BCI_net score. Cohort-level log HR and standard error were extracted from the age-and-stage-adjusted Cox model. Inverse-variance fixed-effect pooling was the primary analysis. DerSimonian–Laird random-effect, Cochran’s *Q* and *I*^2^ were reported as sensitivity diagnostics.

### 4.8 Spatial back-projection

Module z-means were computed at the spot level inside each section, with within-section standardization. Module relationships and high-conductance enrichment were quantified by Spearman correlation and by comparing module scores in the top 15% conductance-sum spots versus the rest using the Mann–Whitney *U* test. Visualizations included four-panel maps of conductance, aggressive, differentiation and BCI_net, and an additive RGB overlay (orange = aggressive, cyan = differentiation, white = co-occurrence) with 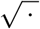 gamma scaling.

### 4.9 PDAC zero-shot validation

Five treatment-naive PDAC Visium sections from GSE274103 were downloaded as sample-level Space Ranger outputs and standardized into the same `filtered_feature_bc_matrix.h5` plus `spatial/` structure used by the OSCC pipeline. No genes or weights were re-estimated in PDAC. Conductance sum, isolation index, edge distance, STC1 + FOS stress, and partial rank correlations were computed as above. The OSCC aggressive module was projected directly; for the structural comparator, a PDAC epithelial/ductal proxy was defined from *KRT8, KRT18, KRT19, EPCAM, MUC1, TACSTD2, CLDN4*, and *KRT7*.

## Data Availability

All original datasets analyzed in this study are publicly available. The spatial transcriptomics datasets can be accessed in the NCBI Gene Expression Omnibus (GEO) under accession numbers GSE208253 and GSE274103. The bulk microarray cohort is available under GEO accession GSE65858. The TCGA-HNSC public cohort data are available via the UCSC Xena data portal. All processed metrics, gene signatures, and intermediate results generated during this study are included in this manuscript and its supplementary information files. Custom Python scripts used for spatial graph construction, directional deconvolution, and statistical modeling are available from the corresponding author upon reasonable request.

https://www.ncbi.nlm.nih.gov/geo/query/acc.cgi?acc=GSE208253

https://www.ncbi.nlm.nih.gov/geo/query/acc.cgi?acc=GSE65858

https://www.ncbi.nlm.nih.gov/geo/query/acc.cgi?acc=GSE274103

https://xenabrowser.net/datapages/?cohort=TCGA%20Head%20and%20Neck%20Cancer%20(HNSC)

## Data and code availability

The 12 spatial sections originate from GEO accession `GSE208253` [Arora et al., 2023]. TCGA-HNSC HiSeqV2 RNA-seq, survival and clinical data were obtained from UCSC Xena. GSE65858 series matrix and GPL10558 platform annotation were obtained from NCBI GEO [Wichmann et al., 2015]. The PDAC validation sections originate from `GSE274103`. All processing scripts (graph construction, diagnostics, ablation, signature extraction, TCGA survival, GSE65858 parsing, mini meta-analysis, spatial back-projection, PDAC zero-shot validation, competing covariate benchmarking and graph sensitivity analysis) are included in the manuscript package and listed in the corresponding methods sections.

## Author contributions

K.T. conceived and supervised the study, designed the analytical framework, and wrote the manuscript. Y.H. and M.C. performed the spatial graph construction, signature extraction, survival analysis, and pan-cancer validation, and contributed to manuscript drafting. All authors discussed the results, contributed to figure preparation, and approved the final version of the manuscript.

## Acknowledgements

The authors thank colleagues for discussion and the original data submitters for making the spatial and bulk cohorts publicly available.

## Supplementary Figures

**Figure S1:**
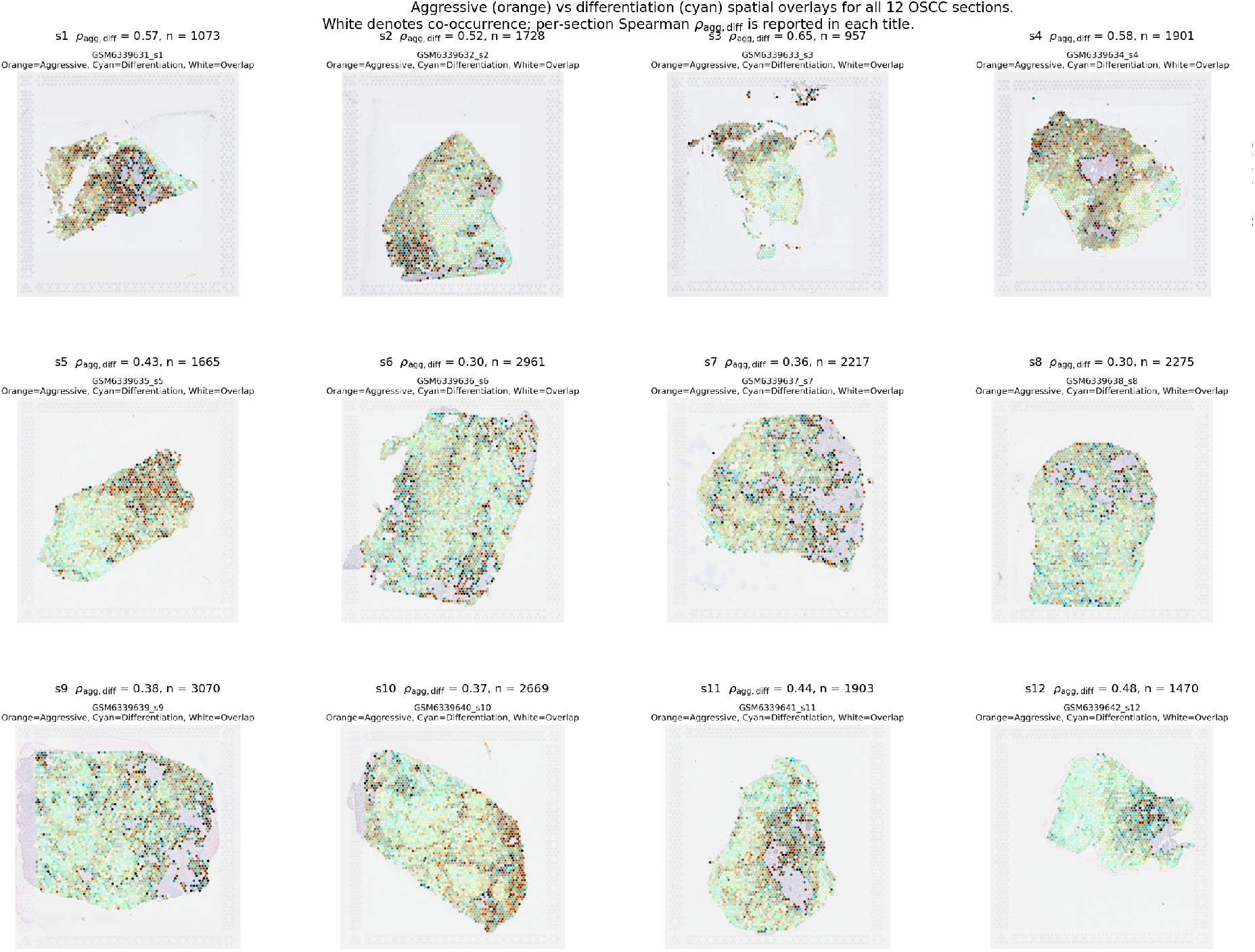
Aggressive (orange) versus differentiation (cyan) spatial overlays for all 12 OSCC Visium sections. White voxels denote spots where both modules are simultaneously elevated. Per-section Spearman *ρ*_agg,diff_ and the number of in-tissue spots are reported in each panel title. All 12 sections show positive co-localization (*ρ* range 0.30–0.65), supporting the microscopic mechanism described in Figure 3.

**Figure S2:**
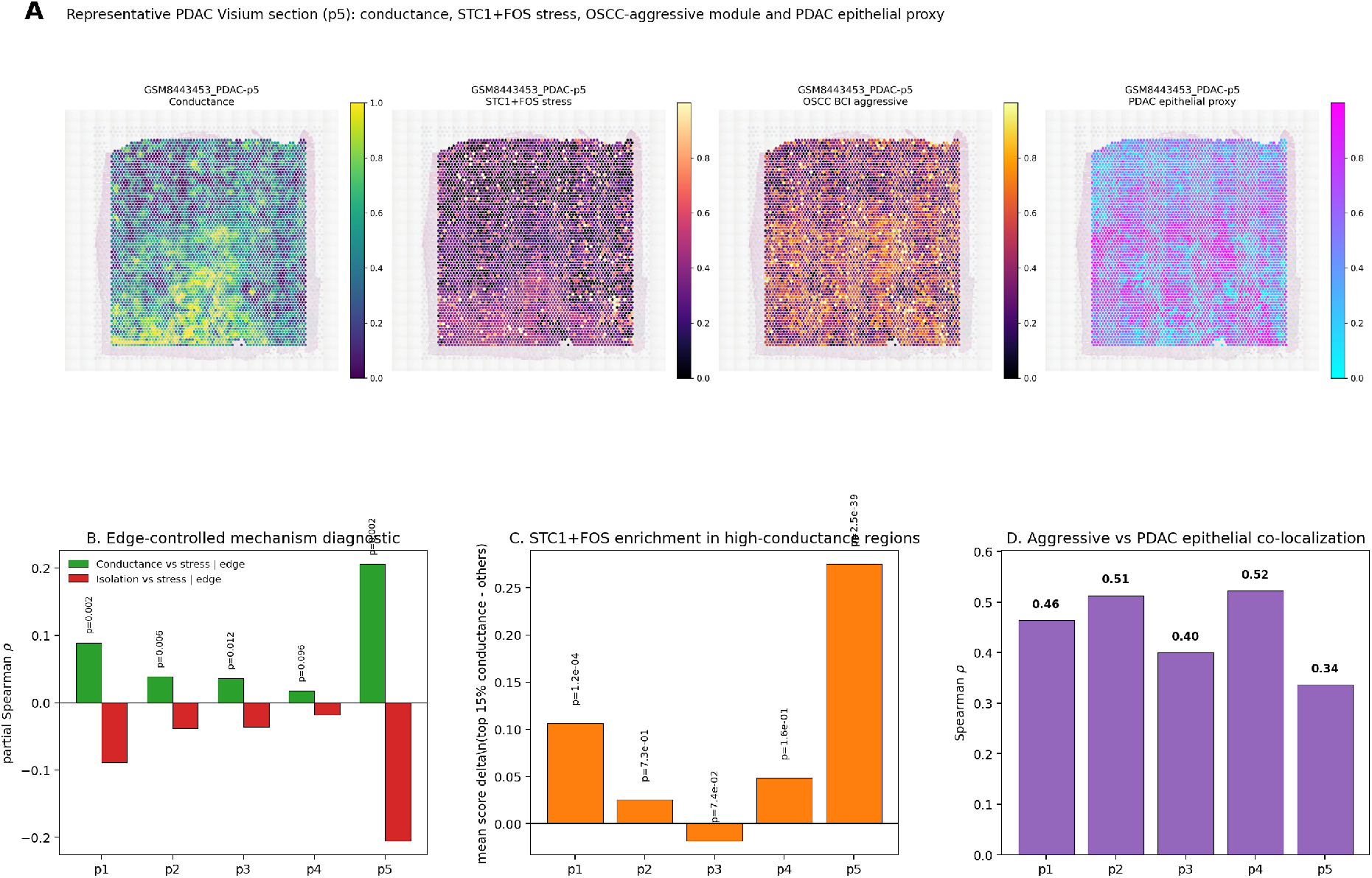
Zero-shot PDAC validation of conductance-centered stress organization. Five treatment-naive PDAC Visium sections from GSE274103 were processed without retraining. The OSCC-derived conductance graph and module projection pipeline was applied directly, with a PDAC epithelial/ductal proxy used as the tissue-structure comparator. Conductance– stress partial correlations were positive in all five sections after edge-distance control, isolation– stress partial correlations were negative in all five sections, and OSCC-aggressive versus PDAC-epithelial module co-localization was positive in all sections.

**Figure S3:**
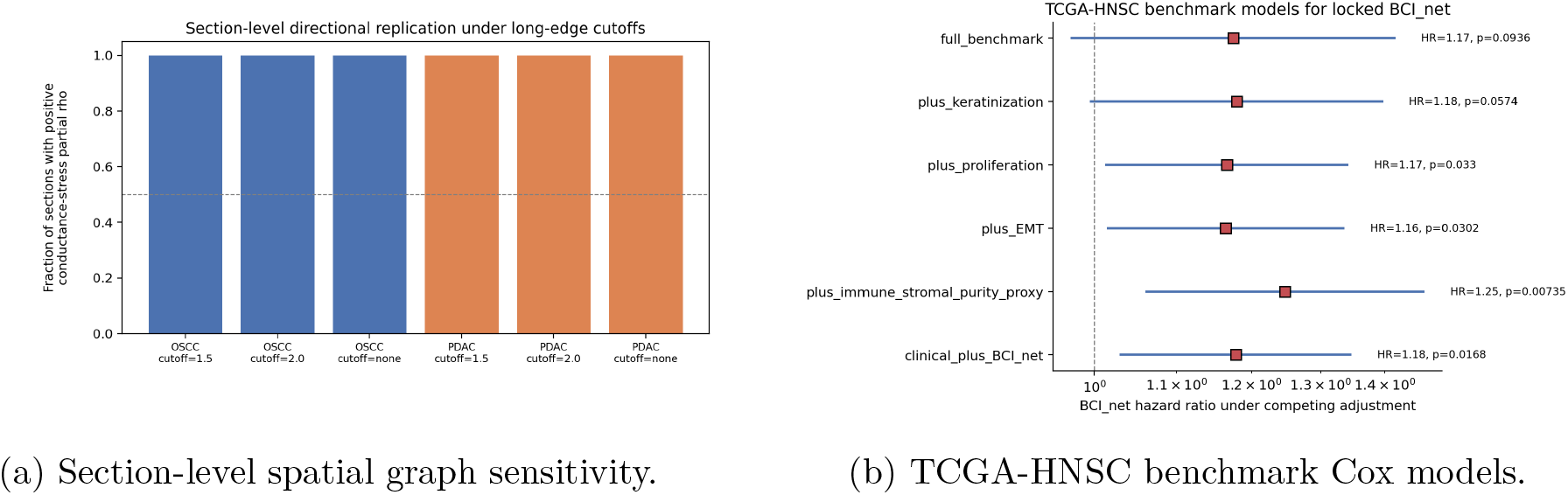
Robustness and sensitivity analyses. **(A)** Fraction of OSCC and PDAC sections preserving positive conductance–stress partial correlation under the primary graph parameters and long-edge cutoff settings. **(B)** Hazard ratio for the locked BCI_net term under competing TCGA-HNSC adjustment models that include clinical variables and transcriptomic proxy covariates for tissue composition, EMT, proliferation and keratinization.

## References

Michael Levin. Molecular bioelectricity: how endogenous voltage potentials control cell behavior and instruct pattern regulation in vivo. Molecular Biology of the Cell, 25(24):3835–3850, 2014. doi: 10.1091/mbc.E13-12-0708.

Michael Levin. Bioelectric signaling: Reprogrammable circuits underlying embryogenesis, regeneration, and cancer. Cell, 184(8):1971–1989, 2021. doi: 10.1016/j.cell.2021.02.034.

Dany Spencer Adams, Joan M. Lemire, Ryan H. Kramer, and Michael Levin. Bioelectric signalling via potassium channels: a mechanism for craniofacial dysmorphogenesis in kcnj2-associated andersen-tawil syndrome. The Journal of Physiology, 594(12):3245–3270, 2017. doi: 10.1113/JP272910.

Trond Aasen, Marc Mesnil, Christian C. Naus, Paul D. Lampe, and Dale W. Laird. Gap junctions and cancer: communicating for 50 years. Nature Reviews Cancer, 16:775–788, 2016. doi: 10.1038/nrc.2016.105.

Gregg L. Semenza. Hypoxia-inducible factors in physiology and medicine. Cell, 148(3):399–408, 2012. doi: 10.1016/j.cell.2012.01.021.

Hyuk-Joon Lee, Heekyung J. Cha, Junho Lim, Sun-Hyun Jeong, Sung-Hyun Park, Cheol-Hee Lee, Jung-Yoon Han, Hyun-Soo Lee, Yeon-Kyun Kim, and Sang-Ho Lee. Cryo-em structures of human cx43 reveal ph-dependent gating mechanism. Nature, 617:839–845, 2023. doi: 10.1038/s41586-023-06002-x.

Patrik L. Ståhl, Fredrik Salmén, Sanja Vickovic, Anna Lundmark, Jose Fernández Navarro, Jens Magnusson, Stefania Giacomello, Michaela Asp, Jakub O. Westholm, Mikael Huss, et al. Visualization and analysis of gene expression in tissue sections by spatial transcriptomics. Science, 353(6294):78–82, 2016. doi: 10.1126/science.aaf2403.

Jeffrey R. Mătt, Emma Lundberg, and Holger Heyn. The emerging landscape of spa-tial profiling technologies. Nature Reviews Genetics, 23:741–759, 2022. doi: 10.1038/s41576-022-00515-3.

Yan Li, Yuting Liu, Cong Wang, Wenjun Xia, Junying Zheng, Jianmin Yang, Bing Liu, Junjie Liu, and Lan Liu. Stanniocalcin 1 (stc1) in cancer: From mechanism to therapeutic potential. Cancer Letters, 500:164–173, 2021. doi: 10.1016/j.canlet.2020.12.008.

Wenjie Sheng, Wei Tan, Bin Wang, Jian Yang, and Zhixin Wang. The cellular and molecular mechanisms of c-fos as a target in cancer therapy. Frontiers in Oncology, 10:583435, 2020. doi: 10.3389/fonc.2020.583435.

Gunnar Wichmann, Maciej Rosolowski, Knut Krohn, Markus Kreuz, Andreas Boehm, Annett Reiche, Universa Müller, Eric Banz, Julia Bertolini, Markus Loíer, et al. The role of hpv rna transcription, immune response-related gene expression and disruptive tp53 mutations in diagnostic and prognostic profiling of head and neck cancer. International Journal of Cancer, 137(12):2846–2857, 2015. doi: 10.1002/ijc.29649.

Rohit Arora, Cunjie Cao, Manoj Kumar, Sarthak Sinha, Ayan Chanda, Rachel McNeil, Divya Samuel, Hardeep K. Arora, T. Wayne Matthews, Shamir Chandarana, et al. Spatial transcriptomics reveals distinct and conserved tumor core and edge architectures that predict survival and targeted therapy response. Nature Communications, 14:5029, 2023. doi: 10.1038/s41467-023-40271-4.

